# Balloon guide catheter effect on first pass and revascularization success

**DOI:** 10.1101/2023.05.11.23289886

**Authors:** Michael D Modzelewski, Sabrina M Perlman, Mark C Duggan, Conrad W Liang

## Abstract

**Background:** The benefit of balloon guide catheter (BGC) use in endovascular thrombectomy (EVT) of patients with acute ischemic stroke remains uncertain. This study assessed the influence of BGC use during EVT on first pass (FP) and revascularization (RV) success in a cohort of stroke patients from a multi-hospital health system.

**Methods:** Patients with anterior circulation large vessel occlusion (LVO) undergoing EVT with stent-retriever or aspiration between 2012 and 2018 at three Kaiser Permanente Southern California (KPSC) region hospitals were identified. A chi-squared test compared the relationship of BGC use with the primary outcomes of FP and RV success using a dichotomized thrombolysis in cerebral infarction (TICI) score of 2b or greater.

**Results:** 218 patients were included. 35 (16%) underwent EVT with BGC. FP success rate did not significantly differ with 37.1% (95% CI 21.5% to 55.1%) FP success in patients that received EVT with BGC and 41.5% (95% CI 34.3% to 49.0%) in patients that received EVT without BGC (p = 0.71). Successful final RV did not significantly differ between the two groups with 85.7% (95% CI 69.7% to 95.2%) final RV success in the EVT with BGC group and 88.5% (95% CI 83.0% to 92.8%) in the EVT without BGC group (p = 0.78). There was no significant difference in FP (p = 0.88) or RV success (p = 0.42) between the BGC (37% FP and 86% RV), non-BGC stent-retriever (42% FP and 92% RV), and aspiration thrombectomy groups (41% FP and 86% RV).

**Conclusion:** There was no observed association between BGC use in EVT of anterior circulation LVO and rates of first pass revascularization or final revascularization.

## Introduction

The development of mechanical thrombectomy devices over the last two decades has transformed treatment guidelines for patients with acute ischemic stroke due to LVO. Endovascular treatment removes clots and rapidly perfuses brain tissue more efficiently than thrombolytic therapy alone^1-3^, which was the principal treatment of acute ischemic stroke prior to FDA approval of the MERCI® retriever in 2004. Today, the most widely used mechanical thrombectomy (MT) techniques use either stent retrievers (SR), contact aspiration thrombectomy (CAT), or a hybrid method that utilizes both techniques^4^.

Although studies show improved reperfusion and better clinical outcomes with MT than thrombolytic treatment alone^5-8^, a theoretical risk of the procedure is the generation of distal emboli due to clot fragmentation upon insertion of guiding microwires and subsequent microcatheters^9-10^, leading to failure of recanalization and increased reperfusion time to ischemic brain tissue. A proposed solution to this complication is the use of a BGC inflated proximal to the clot, thus arresting antegrade flow and minimizing the risk of distal emboli flow. Advocates cite studies that associate BGC use with lower rates of distal emboli formation and better first-pass recanalization rates, thus decreasing time to tissue reperfusion and improving the quality of reperfusion^11-13,17^.

Adoption of BGC use during MTs has not been universal among neurointerventionalists. A 2020 meta-analysis of EVT device use found that less than 50% of MTs are performed with a BGC^20^. Arguments against BGCs include their varying compatibility with certain MT devices, reported vascular complications^16^, and the opinion that the catheters used in MT procedures are enough to partially occlude antegrade flow and adequately reduce the risk of distal emboli^14-15^. In addition, a few studies have contradicted previous findings of BGC EVT superiority and demonstrated no significant difference in clinical or reperfusion outcomes between BGC and non-BGC EVT^18-19^, though these studies compared BGC SR-MT with combination SR-CAT. Notably, the most recent and largest study to demonstrate non-superiority of BGCs is the 2022 analysis of the ASSIST registry which assessed the outcomes of 1,300 thrombectomies and found no difference in FP or final RV scores between BGC EVT and non-BGC EVT groups^21^.

Currently, neurointerventionalists remain divided on BGC utility due to contradictory study results and competing provider preferences. To assess BGC influence and better inform treatment guidelines, we conducted a retrospective analysis of KPSC patients to investigate differences in revascularization and clinical outcomes between MTs performed with a BGC versus those performed without a BGC.

## Methods

Institutional review board approval was obtained for a retrospective chart review. The inclusion criteria for this study were defined as patients diagnosed with anterior circulation large vessel occlusion that underwent endovascular clot retrieval using a stent-retriever or aspiration device between the years 2012-2018 at three thrombectomy-capable KPSC medical centers. Potential patients were identified from hospital stroke coordinator and interventionalist case logs.

Eligibility according to inclusion and exclusion criteria was confirmed by manual chart review. Chart review of the patient medical record was conducted to determine which patients were treated with BGC. Brand, model, and size of the BGC were not assessed for the purposes of this study. Patient characteristics collected from chart review were the initial National Institute of Health Stroke Scale (NIHSS) score, administration of tissue plasminogen activator (tPA) prior to EVT, patient demographics (age, race, sex), and select medical history limited to conditions associated with higher stroke risk and/or complications: diabetes mellitus, hypertension, atrial fibrillation (AF), previous myocardial infarction, and history of or current tobacco use.

The primary outcome of the study compared FP RV success and final RV outcome in the BGC versus non-BGC groups, data for which was obtained from the previous study’s data set. From this data set, our study also collected information on the MT device used for first RV attempt (aspiration, Capture^™^, Solitaire^™^ SR or Trevo® SR). FP success was documented during the procedure (y/n) and final RV outcome is based on the provider determined TICI score (0, 1, 2a, 2b, 2c, 3), with 2b or greater representing successful revascularization.

Secondary outcome variables were obtained through manual chart review. Patient clinical outcomes were assessed by comparing final documented NIHSS score prior to discharge (low: 0-14, intermediate: 15-28, high: 29-42) and discharge disposition (acute care or rehab facility, skilled nursing facility (SNF), home, home with hospice, deceased) between the BGC and non-BGC groups.

The effect of BGC use on the primary and secondary outcome variables was measured using a chi-squared test. Multivariable logistic regression assessed the relationship between BGC use and final RV success with the following confounder/effect modifier variables: age, gender, select medical history, initial NIHSS score severity and tPA administration.

## Results

### Study Cohort

218 patients that met inclusion criteria were identified, of which 35 (16.1%) underwent EVT with BGC. The median patient age was 72 years and 94 of the patients (43.1%) were female. Comparison of patient characteristics between the BGC and non-BGC groups demonstrated no significant difference in age (p = 0.662), race (p = 0.706), sex (p = 1), initial NIHSS score (p = 0.619), tPA administration prior to EVT (p = 0.384), or relevant medical history. Relevant medical history and patient demographic data are shown in Table 1 below.

**Table 1.** Patient demographics and past medical history; provider and medical center data.

|  | **No BGC (n= 183)** | **BGC (n= 35)** | **P-value** |
| --- | --- | --- | --- |
| **Age** | 69.9 +/- 13.5 | 68.5 +/- 17.9 | 0.66 |
| **Sex** |  |  | 1.00 |
| Female | 79 (43.2%) | 15 (42.9%) |  |
| Male | 104 (56.8%) | 20 (57.1%) |  |
| **Race** |  |  | 0.71 |
| Asian | 23 (12.6%) | 3 (8.6%) |  |
| Black | 34 (18.6%) | 11 (31.4%) |  |
| Hispanic | 56 (30.6%) | 8 (22.9%) |  |
| White | 60 (32.8%) | 12 (34.3%) |  |
| Other | 5 (0.01%) | 0 (0.0%) |  |
| Missing | 5 (2.7%) | 1 (2.9%) |  |
| **Presenting NIHSS** |  |  | 0.64 |
| 1-14 | 65 (35.5%) | 11 (31.4%) |  |
| 15-28 | 113 (61.7%) | 22 (62.9%) |  |
| 29-39 | 5 (2.7%) | 2 (5.7%) |  |
| **tPA given** | 92 (50.3%) | 21 (60.0%) | 0.38 |
| **Serum glucose** | 141 +/- 57.1 | 131 +/- 37.2 | 0.25 |
| **Medical history** |  |  |  |
| Hypertension | 143 (78.1%) | 26 (74.3%) | 0.78 |
| Diabetes mellitus | 63 (34.4%) | 11 (31.4%) | 0.85 |
| Atrial fibrillation | 88 (48.1%) | 14 (40.0%) | 0.53 |
| Myocardial infarction | 30 (16.4%) | 4 (11.4%) | 0.68 |
| Tobacco use | 75 (41.0%) | 16 (45.7%) | 0.74 |
| **Site** |  |  | 0.17 |
| Anaheim | 9 (4.9%) | 0 (0.0%) |  |
| Fontana | 24 (13.1%) | 2 (5.7%) |  |
| Los Angeles | 150 (82.0%) | 33 (94.3%) |  |
| **Provider** |  |  | <0.001 |
| Physician 1 | 98 (53.6%) | 7 (20.0%) |  |
| Physician 2 | 52 (28.4%) | 25 (71.4%) |  |
| Physician 3 | 0 (0.0%) | 1 (2.9%) |  |
| Physician 4 | 16 (8.7%) | 0 (0.0%) |  |
| Physician 5 | 2 (1.1%) | 0 (0.0%) |  |
| Physician 6 | 7 (3.8%) | 0 (0.0%) |  |
| Physician 7 | 8 (4.4%) | 2 (5.7%) |  |

### Procedure data

The majority of EVT procedures were performed at Kaiser Permanente (KP) Los Angeles Medical Center (183, 83.9%) followed by KP Fontana Medical Center (26, 11.9%) and KP Anaheim Medical Center (9, 4.1%), with no significant difference in prevalence of BGC use between the medical centers (p = 0.165). Among the seven providers performing EVT, there was a significant difference (p < 0.001) in BGC use with two providers accounting for 91.4% of all BGC assisted EVT, both of whom operated exclusively at KP Los Angeles Medical Center. Detailed provider and medical center data are shown in Table 1 above. All EVT performed with BGC used stent retrievers as the first thrombectomy device in the procedure, either the Trevo® SR (19/35, 54.3%) or the Solitaire^™^ SR (16/35, 45.7%). First pass devices utilized in EVT without BGC were aspiration (96/183, 52.5%), Solitaire^™^ SR (52/183, 28.4%) and Trevo® SR (34/183, 18.6%), and Capture^™^ device (1/183, 0.5%).

### Effect of BGC on FP and RV success

FP success rate did not significantly differ (p= 0.71) between the BGC group (37.1%, 95% CI 21.5% to 55.1%) and non-BGC group (41.5%, 95% CI 34.3% to 49.0%), shown in Figure 1.

**Figure 1.**
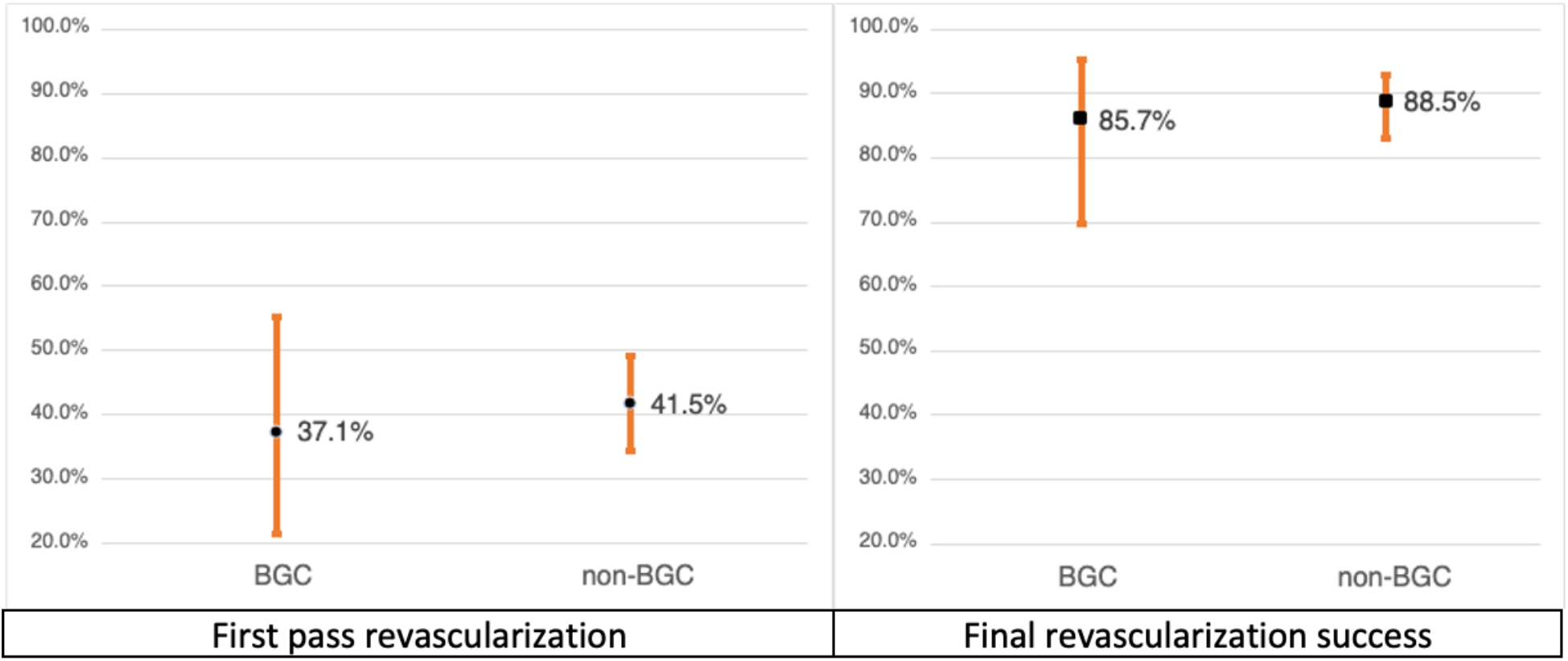
Effect of BGC on FP and RV success.

Successful final RV, measured as a TICI score of 2b or greater, did not significantly differ (p = 0.78) between the BGC group (85.7%, 95% CI 69.7% to 95.2%) and non-BGC group (88.5%, 95% CI 83.0% to 92.8%), shown in Figure 1. Detailed reperfusion outcomes are shown in Table 2.

**Table 2.** Reperfusion and clinical outcome data.

|  | **No BGC (n= 183)** | **BGC (n= 35)** | **P-value** |
| --- | --- | --- | --- |
| **First pass success** | 76 (41.5%) | 13 (37.1%) | 0.71 |
| **Final revascularization success (TICI 2b or greater)** | 162 (88.5%) | 30 (85.7%) | 0.78 |
| **TICI score** |  |  | 0.49 |
| 0 | 4 (2.2%) | 1 (2.9%) |  |
| 1 | 2 (1.1%) | 2 (5.7%) |  |
| 2a | 14 (7.7%) | 2 (5.7%) |  |
| 2b | 77 (42.1%) | 14 (40.0%) |  |
| 2c | 19 (10.4%) | 2 (5.7%) |  |
| 3 | 67 (36.6%) | 14 (40.0%) |  |
| **Final NIHSS** |  |  | 0.24 |
| 0-14 | 126 (68.9%) | 18 (51.4%) |  |
| 15-28 | 26 (14.2%) | 8 (22.9%) |  |
| 29-42 | 1 (0.5%) | 0 (0.0%) |  |
| Missing | 30 (16.4%) | 9 (25.7%) |  |
| **Discharge Disposition** |  |  | 0.22 |
| Acute care or rehab facility | 21 (11.5%) | 2 (5.7%) |  |
| Skilled nursing facility | 52 (28.4%) | 14 (40.0%) |  |
| Home | 85 (46.4%) | 10 (28.6%) |  |
| Home with hospice | 1 (0.5%) | 1 (2.9%) |  |
| Deceased | 24 (13.1%) | 8 (22.9%) |  |
| **Length of Stay** |  |  | 0.58 |
| Mean (SD) | 7.71 (6.75) | 8.52 (7.08) |  |
| Median [min, max] | 5.00 [1.00, 37.0] | 6.00 [1.00, 36.0] |  |
| Deceased | 24 (13.1%) | 8 (22.9%) |  |

There was no significant difference in FP (p = 0.88) or RV success (p = 0.42) between the BGC (37% FP and 86% RV), non-BGC SR-MT (42% FP and 92% RV), and non-BGC CAT (41% FP and 86% RV) groups.

### Effect of BGC on clinical outcomes

An NIHSS score in the post-procedure period prior to discharge was available for 153 patients (83.6%) in the non-BGC group and 26 patients (74.3%) in the BGC group. There was no significant difference in final NIHSS score (p = 0.24) between the BGC group (low: 51.4%, intermediate: 22.9%, high: 0.0%) and non-BGC group (low: 68.9%, intermediate: 14.2%, high: 0.5%). Discharge disposition data were available for all 218 patients and demonstrated no significant difference (p = 0.22) between the groups. In the non-BGC group, 21 (11.5%) went to acute care or rehab facilities, 85 (46.4%) were discharged home, 1 (0.5%) was discharged home with hospice, 52 (28.4%) went to a SNF and 24 (13.1%) died prior to discharge. In the BGC group, 2 (5.7%) went to acute care or rehab facilities, 10 (28.6%) were discharged home, 1 (2.9%) was discharged home with hospice, 14 (40.0%) went to a SNF and 8 (22.9%) died prior to discharge. There was no significant difference in length of hospital stay (0.583) between the groups. Clinical outcomes are shown in Table 2.

### Multivariable regression analysis

In regression analysis, there was no significant difference in final RV success between the BGC and non-BGC groups. Age, gender, administration of tPA and high NIHSS score were not significantly associated with improved final RV success. Among relevant past medical history, only a history of AF (p = 0.0368) was demonstrated to have a significant effect on final RV. Detailed regression analysis data are shown in Table 4.

**Table 4.** Multivariable regression analysis.

|  | OR | P-value | 95% CI |
| --- | --- | --- | --- |
| **Final revascularization success (TICI 2b or greater)** |  |  |  |
| Age | 0.987 | 0.532 | 0.947 – 1.03 |
| Gender | 0.870 | 0.793 | 0.296 – 2.44 |
| NIHSS score 15-28 | 0.488 | 0.201 | 0.153 – 1.41 |
| NIHSS score 29-39 | 0.192 | 0.238 | 0.0137 – 5.10 |
| tPA administered | 2.56 | 0.0768 | 0.921 – 7.59 |
| Hypertension | 1.61 | 0.454 | 0.442 – 5.54 |
| Diabetes mellitus | 2.47 | 0.239 | 0.619 – 13.7 |
| Atrial fibrillation | 3.81 | 0.0368 | 1.14 – 14.7 |
| Myocardial infarction | 1.94 | 0.383 | 0.491 – 10.4 |
| Smoking | 1.05 | 0.928 | 0.378 – 2.98 |

## Discussion

Our study found no statistically significant difference in FP or final RV success between BGC MT versus non-BGC MT. In addition, there was no statistically significant difference in final NIHSS score and discharge disposition, the two clinical outcomes assessed in this study. Our findings do not support conclusions of previous studies that demonstrated superior outcomes associated with BGC use; for example, Velasco et al. However, our findings add weight to the argument that BGC may not be a critical component of effective MT, as does the data from the 2022 ASSIST registry. These findings do not advocate for the elimination of BGC use altogether but instead demonstrate that the decision to forego BGC use may not significantly alter patient outcomes and can remain at the discretion of the neurointerventionalist. Notably, our multivariable regression analysis demonstrated statistically significant final RV in patients with AF, suggesting occlusions of cardioembolic origin may benefit from BGC use though further research is required to establish this link more conclusively. The role of MT as a widely used yet quickly evolving stroke therapy warrants continuous research on the varying devices and techniques of providers to ensure patients receive the most effective and safe care. No study to date has done an in-depth assessment of the low BGC adoption rate despite research that demonstrates the improved effect of BGC on outcomes. Future research that surveys neurointerventionalists to assess why one MT technique is preferred to another may better explain this discrepancy and guide improvements in BGC devices. In addition, future work that incorporates more thrombectomy cases would yield a larger dataset and may provide more conclusive results on the effects of BGC in MT.

### Limitations

The study has several limitations. First, the number of patients in the BGC group compared to the non-BGC group (35 vs 183) resulted in a lower than anticipated power, limiting the strength of our conclusion that no difference exists between the two groups. Second, tandem occlusions and clot burden, two factors which studies have shown to influence RV and clinical outcomes^22-24^, were not considered in the study analysis thus possibly influencing RV success rates in both groups and limiting the accuracy of our data. Third, the certainty of our secondary outcome conclusion that no difference in clinical outcomes exists between the groups was weakened due to the lack of a long-term outcome metric, such as 90-day mRS score, and inconsistent final NIHSS scores. Clinical outcome data collected prior to discharge do not examine patient functionality in the long stroke recovery period, which can be influenced heavily by the efficacy of EVT reperfusion^25^. In addition, inconsistent final NIHSS scores led to non-standardized comparison between patients with wide variations between the time from EVT procedure to the final documented NIHSS score due to non-standardized assessment intervals, poor provider documentation and varying patient hospital stay lengths. Fourth, not including BGC brand, model, size, and deployment site in each procedure limited our analysis of confounding variables, though this information was not reported in procedure notes. BGC specifications, particularly the distance between BGC deployment and thrombus, have been shown to influence reperfusion outcomes^26^ and may have affected RV success in the BGC group. Further, the timeframe of our study aligns with a period of significant evolution in BGC technology with the development of the CELLO^™^ and FlowGate® BGCs. Comparing the prevalence and outcomes of these newer generation BGCs to the earlier Merci® BGC may have provided a more accurate analysis of our data set. Lastly, BGC use is not randomized, but instead used at the discretion of the physician with various influencing factors not assessed in this study such as provider experience or preference, vessel tortuosity or access difficulty, and BGC cost or hospital supply. These non-random factors, particularly provider experience and vessel characteristics, may have influenced reperfusion outcomes thus limiting confounder analysis and the certainty of our conclusion that no difference exists.

## Conclusion

In this multi-center cohort, BGC use in MT of anterior circulation LVO was not associated with higher rates of first pass revascularization or final revascularization success when compared with MT performed without a BGC. The findings of this study demonstrate that BGC use does not significantly influence patient reperfusion outcomes and suggest BGC use remain at the discretion of the individual neurointerventionalist. The small number of patients in the BGC group relative to the non-BGC group affects the strength of our results though a larger dataset that utilizes inter-regional KP thrombectomy data may provide a more conclusive result.

## Data Availability

The data that support the findings of this study are available on request from the corresponding author, MM. The data are not publicly available due to their containing information that could compromise the privacy of research participants.

## List of abbreviations

BGC: balloon guide catheter
EVT: endovascular thrombectomy
FP: first pass
RV: revascularization
LVO: large vessel occlusion
KPSC: Kaiser Permanente Southern California
TICI: thrombolysis in cerebral infarction
MT: mechanical thrombectomy
SR: stent retriever
CAT: contact aspiration thrombectomy
NIHSS: National Institute of Health Stroke Scale
tPA: tissue plasminogen activator
AF: atrial fibrillation
SNF: skilled nursing facility
KP: Kaiser Permanente

